# Establishing emergency bronchoscopy in Timor-Leste: a health care needs assessment-driven novel model of training

**DOI:** 10.1101/2025.09.11.25332169

**Authors:** Helio Guterres, Celia Santos, Agustinha da Silva Soares, Dianne Jones, Chris Kiely, Chris Hair, Finlay Macrae, Daniel P Steinfort

## Abstract

**Background:** Bronchoscopy is an integral component of the practice of respiratory medicine. Multiple challenges to establishing and maintaining a service in low income countries (LIC) are recognized, with establishing and maintaining bronchoscopic competence being a key barrier to implementation. A critical gap in care at the leading hospital in Dili, Timor-Leste, was identified following multiple adverse outcomes due to lack of bronchoscopy for management of foreign body aspiration (FBA) in 2023, including two deaths.

**Methods:** A healthcare needs assessment (HCNA) was completed to identify setting-specific requirements, and a novel training program designed to rapidly upskill local clinicians for performance of emergency FBA retrieval.

**Results:** HCNA revealed heightened risks (no negative pressure ventilation, limited anaesthetic staff capacity) and reduced diagnostic utility (lower lung cancer rates, no treatment for advanced cancer or TB strictures), but noted significant infrastructure and workforce capacity for bronchoscopy due to the endoscopy training program established by the Australian and New Zealand Gastroenterology International Training Association (ANZGITA). A novel training program, comprising short intensive skills training and ongoing remote mentorship (including ‘just-in-time procedural support) was implemented to rapidly upskill local HNGV clinicians. In the sixteen months following bronchoscopic training, three paediatric patients successfully underwent bronchoscopic removal of FBA without complications.

**Conclusion:** This model demonstrates the value of intensive training and remote support for successful establishment of bronchoscopy for targeted indications, and offers a potential framework for other LICs to establish essential emergency bronchoscopy services despite resource limitations and low procedural volumes

---

Bronchoscopy is an integral component of practice of respiratory medicine in many countries, though services are often limited in low-income countries (LIC) due to limitations in the human and equipment factors critical for success and sustainability. These challenges are similar to those that are well documented in establishing other endoscopic services in low resource settings, including upper and lower gastrointestinal endoscopy and cystoscopy.(1)

Hospital Nacional Guido Valadares (HNGV) is the main hospital for Dili (population 222,000) and serves as the referral centre for all of Timor-Leste (population 1.38m). The HNGV endoscopy service was developed in partnership with the Australian and New Zealand Gastroenterology International Training Association (ANZGITA). Since 2016, ANZGITA has guided development of a robust, sustainable, safe endoscopy unit in Dili in collaboration with the Ministry for Health, Timor-Leste.

In 2023, over 1,100 upper gastrointestinal endoscopic procedures and 200 colonoscopy procedures were performed, with no significant complications observed. In the same period, five patients presented to HNGV emergency department with inhaled airway foreign bodies (FBA). With no bronchoscopy service available, patients were managed via air evacuation to neighbouring countries for surgical retrieval of FBA via thoracotomy and bronchotomy. Unfortunately, two patients died prior to evacuation.

Medical leadership at HNGV contacted ANZGITA for support in establishment of a bronchoscopy service to complement their existing endoscopic service. ANZGITA facilitated an in-country focused health care needs assessment (HCNA) to assess the utility of bronchoscopy at HNGV, utilizing established frameworks.(2) In-country review included attendance by an experienced endoscopy nurse (DJ) and Interventional Pulmonologist (DPS) as well as consultation with lead clinicians at HNGV to inform needs assessment.

Key observations of HCNA included:

1. Heightened setting-specific risks
  - absence of negative-pressure ventilation in endoscopy suites
  - absence of anaesthetic support for conscious sedation procedures.

1. Reduced diagnostic utility
  - Lower rates lung cancer (per population)
  - no treatment options available for patients with locally advanced/metastatic lung cancer, or post-tuberculous stricturing

1. Service needs assessment
  - Existing factors/competencies present
    - equipment factors (e.g. video processors, sterilization equipment)
    - facility factors (ergonomic space for theatre and recovery, including monitoring)
  - workforce mapping
    - experienced endoscopic nursing/tech staff
    - highly motivated senior medical staff
    - established endoscopy/recovery workflow

With uncertain utility of bronchoscopy and the potential for harm, establishment of standard bronchoscopy service was considered not viable.

In contrast, service needs assessment identified presence of critical resources to support a bronchoscopy service, and clinicians at HNGV remained highly motivated to establish a bronchoscopy service for management of future airway emergencies.

Recognizing the challenge of developing and maintaining competency in bronchoscopy, we sought to establish the necessary skillset within HNGV clinicians via a novel model of bronchoscopic training using a program developed specifically for HNGV, Dili, leveraging existing infrastructure and workforce capacity established at HNGV by the ANZGITA program.

An Internal Medicine physician (HG) and experienced endoscopy nurse (AS) from Timor-Leste were supported with a 4-day observership at tertiary bronchoscopy centres in Australia. HG was also provided with a low-fidelity 3D-printed airway model for unstructured simulation training, as previously described,(3, 4) and attended a two-day bronchoscopy focused skills workshop. On return to Dili, a remote support program was established via WhatsApp to provide case/imaging review from an Interventional Pulmonologist for case formulation and, following confirmation of indication for bronchoscopy, ‘just-in-time’ procedure planning.(5)

From 23^rd^ February 2024 to 23^rd^ June 2025, four patients presented to HNGV with FBA (age range 2-12years). Flexible bronchoscopy was performed via laryngeal mask airway under general anaesthesia via laryngeal mask airway using a 5.5mm diameter bronchoscope (Olympus BF-Q180, Olympus; Tokyo, Japan). Bronchoscopic retrieval of FBA was unsuccessful in one 6.5kg boy, due to bronchoscope size precluding introduction into the airways. The child was evacuated for thoracotomy retrieval of FBA. Bronchoscopic FBA retrieval was successful in three cases; a boy with aspirated stone in distal trachea (figure 1a,b), a boy with an aspirated needle (figure 1c,d), and a boy with an aspirated coffee bean occluding the proximal right main bronchus. All patients were discharged one day following bronchoscopy without any complication.

**Figure 1:**
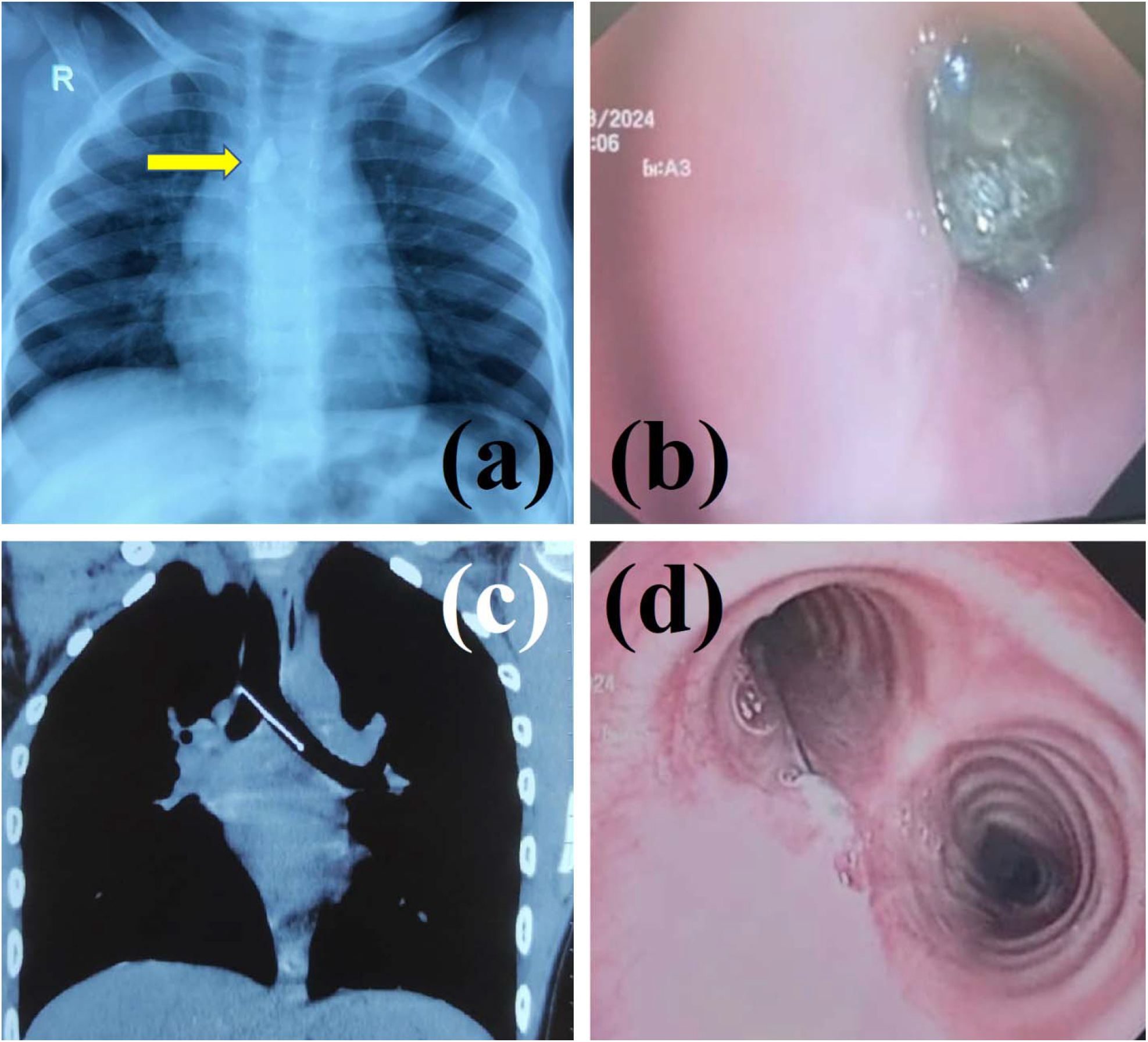
Following presentation of a young boy to emergency room with a history of choking & cyanosis, CXR (a) demonstrated radio-opaque foreign body in distal trachea (arrow). Bronchoscopy (b) demonstrated impaction of stone in distal trachea, with near complete luminal occlusion. A high-school aged boy presented with a clear history of aspirating a needle. (c) CT chest (c) demonstrated aspirated pin at main carina, extending into proximal left main bronchus. Bronchoscopy (d) confirming pin resting on posterior wall of proximal left main bronchus.

Foreign body aspiration (FBA) is an airway emergency, and remains one of the commonest causes of death in young children.(6) Surgical thoracotomy and bronchotomy is required where FB is unable to be retrieved bronchoscopically,(7) though is associated with significant morbidity and mortality, as well as high resource utilization. As the NHGV experience in 2023 demonstrates, there remains a critical role for bronchoscopy in LICs.

Our novel model of intensive skills training combined with remote mentoring, leveraging the existing endoscopic competencies (equipment/facility factors, nursing expertise) established by ANZGITA, proved highly effective in establishing an effective emergency bronchoscopy service, as evidenced by prevention of death or major morbidity in three patients with FBA. This lifesaving intervention aligns with the The *Lancet* Commission on Global Surgery ‘s (8) goal of timely essential surgical care and ensuring universal access to safe and affordable procedural care.

Concerns regarding establishment of bronchoscopy in LIC include funding for equipment, acquisition and maintenance of bronchoscopic competency as well as service and maintenance costs and availability of equipment.(11)(16) Lack of training is another major barrier.(16) Bronchoscopy training programs in high-resource settings frequently combine procedural simulation, with supervised stepwise escalation in procedural complexity and risk and while competence assessment is increasingly valued, most professional societies remain focused on procedural volume (over multiple years) as a precondition of competence. Due to low caseloads, absence of existing expertise and competing workloads, it is difficult for LIC clinicians to attend a traditional 6-12 month fellowship in a high-volume centre to develop competency. An alternative model of training is required for LIC.

Our model of physician-performed bronchoscopy with a short intensive fellowship including ongoing remote mentoring support, effectively addresses barriers to competence and may prove a model to similar resource-limited settings to be able to establish the key competency of bronchoscopy for FBA retrieval. Use of 3D airway models for simulation addresses the inevitable decay in dexterity skills resulting from low procedural volume.(4) Our model leverages the successful endoscopic service established by ANZGITA, including equipment factors (e.g. video processors, sterilization equipment), facility factors (ergonomic space for theatre and recovery, including monitoring), procedural clinician and nursing competencies (including case planning and recovery.)

Remote mentorship is an effective tool in gastroscopy and colonoscopy training, and has been proven to overcome isolation faced by clinicians in low resource environments. Remote mentoring by video (eg zoom/teams/WhatsApp etc) may be a way forward for real-time (or just-in-time (5, 20)) support during emergency cases in bronchoscopy, as demonstrated previously in gastrointestinal endoscopy.(21, 22)

In conclusion, current models of bronchoscopic training and competence assessment are not suitable models for low-income countries. We demonstrate a novel model – developed in partnership with clinicians at HNGV – for successful establishment of bronchoscopy in LICs, where indications for bronchoscopy are rare, comprising short intensive skills training and supported with simulation, remote mentoring and ‘just-in-time’ case review, leveraging existing institutional endoscopy skills and resources. The value of bronchoscopy for management of airway emergencies is high, as demonstrated by successful potentially life-saving bronchoscopic retrieval of FBA. This model of targeted service improvement with ongoing mentorship may provide a model for other LICs to develop necessary services for their patient populations

## Data Availability

All data produced in the present work are contained in the manuscript

## Data sharing

No data will be made available

